# Immune monitoring of pediatric patients co-infected with *Rickettsia rickettsii* and *Ehrlichia canis*

**DOI:** 10.1101/2022.08.23.22279108

**Authors:** Laura Garcia-Rosales, Angelica Escarcega-Avila, Moises Ramirez-Lopez, Diana Manzanera-Ornelas, Enrique Guevara-Macias, Maribel Vaquera-Arteaga, Carolina Alvarado-Gonzlaez, Blanca Elisa Estrada, Florinda Jimenez-Vega, Luis Donis-Maturano, Gerardo Pavel Espino-Solis

## Abstract

In 2021, 273 Rocky Mountain Spotted Fever cases have been reported nationwide. In Chihuahua City, fourteen samples were obtained from children suspected of rickettsial infection. The analysis of samples collected from January to December 2021 showed a prevalence of 28.5%, 43% and 28.5% for Rickettsia rickettsii, Ehrlichia canis, and both pathogens in coinfection, respectively. The analysis of clinical hematological and biochemistry analytes showed alterations such as 100% of the children coursed with elevated liver enzymes and coagulation times, 64% showed leukocytosis due to neutrophilia, 55% of them had thrombocytopenia, lymphopenia and hypoalbuminemia, and 45% showed normocytic normochromic anemia. Statistically significant differences were obtained in the chemokines IL-8, RANTES, CXCL9/MIG, and CXCL10/IP-10 across the coinfected and control groups; the differences in IP-10 were significant for patients infected by R. rickettsii compared to the control group. Also, significant differences were observed for IL-1β, IL-6, IL-17, IFNγ, and TNFα among the R. rickettsii positive group compared to the control group; on the other hand; the coinfected group exhibited modified levels of IL-6, IL-8, and IL-10 compared with the control group. Finally, significant differences were obtained for CD8 + T lymphocytes subpopulations between positive individuals for R. rickettsii and E. canis.

## Introduction

Tick-borne diseases have public importance from clinical and veterinary perspectives in Mexico, especially the often-neglected Rocky Mountain Spotted Fever (RMSF) caused by the bacteria *Rickettsia rickettsii*, with 273 tick-borne cases reported nationwide in Mexico in 2021(1). The State of Chihuahua reported 76 confirmed cases, of which 59 were caused by RMSF and 17 by another rickettsiosis not identified. This number of cases has pushed this state into the second place of increasing clinical cases reported in Mexico from 2020 to 2021 (1). These tick-borne diseases are responsible for a high lethality in infants, requiring immediate monitoring (2).

Rickettsiosis has been recognized since 1930 in the northern border states of Mexico; from 1985 till the present, these ailments are responsible for outbreaks in Sinaloa, Sonora, Durango, Nuevo León, Coahuila (3,4), and Southeastern states of U.S.A. (5,6). Different research groups reported a comeback of Rickettsiosis was reported in the first years of the 21st century by, with lethality ranging from 20% to 70% [7–10]. Likewise, Rickettsiosis is classified as an emerging disease of public health importance in Mexico, where thousands of cases and hundreds of deaths have been reported in humans (7,8) due to the difficulty of its diagnosis and fatal outcomes when there is no timely intervention in treatment (9,10). Furthermore, it is considered an occupational risk disease (11) and is important for travelers, especially in states bordering the north of Mexico (12). Rickettsial diseases are caused by intracellular bacteria belonging to the Rickettsial order like *R. rickettsii, E. canis*, and *Anaplasma phagocytophilum* (13).

In northern Mexico, these infections are classified as zoonotic vector-borne diseases. The vector *Rhipicephalus sanguineus*, also known as the brown dog tick, has a cosmopolitan distribution and is frequently founded in the peridomestic environment, causing massive infestations as a consequence of its introduction by dogs (14,15). Several bacteria, viruses, and protozoa are transmitted by ticks, but bacteria are responsible for the vast majority of the reported tick-transmitted infections (16,17).

Rickettsial diseases clinical manifestations range from a febrile and unspecific syndrome with temperatures >40°C that progress to a multisystemic organ failure and death if it is not diagnosed and treated in time (18). Patients may also present other unspecific symptoms such as vomiting, nausea, headache, myalgia, arthralgia, and more; consequently, these illnesses are usually confused with other common pathologies in children (19). These ailments may cause various respiratory, gastrointestinal, nervous, and other clinical symptoms (20). Additionally, they are characterized by stimulating an inflammatory response that is reflected in the clinical alterations observed in the acute phase of infection (21); and alterations such as anemia, thrombocytopenia, lymphocytosis, leucopenia, elevated liver enzymes, and coagulation times (prothrombine, and thromboplastin), hipoalbuminemia, hipoproteinemia and others may be observed (22). These clinical signs are associated with the expression and secretion of different proinflammatory mediators (17) and are sometimes potentiated by the presence of other infectious agents in coinfection (23). Coinfections aggravate the clinical signs, complicate the diagnosis, and reduce the time frame for opportune, correct, and specific diagnosis and treatment (24).

Concerning pediatric cases of Rickettsiosis caused by *R. rickettsii*, Martínez-Medina et al., 2005 recorded two pediatric cases in Sonora(25); Leon - Arias et al., 2008, reported eight confirmed cases of Rocky Mountain Spotted Fever (RMSF), which 75% of the patients were infants and 50% of them ended in fatality (26). In a 9-year retrospective study conducted by Gómez-Rivera et al., 2013, at the Sonora State Children’s Hospital, they recorded a total of 116 children considered as probable positive, of which only 86 (74.6%) were confirmed (Gómez et al., 2013). Moreover, in a cross-sectional study of 47 patients who died of RMSF carried out by Delgado-De la Mora et al., 2018, they reported that 49% of affected patients were infants over three years in the state of Sonora (27).

Regarding co-infections, it has been reported co-infection of Dengue virus and *R. rickettsii* in a woman [31] and a co-infection of *R. rickettsii* and *Leptospira spp* in a 12-year-old boy who presented nonspecific signs that rapidly progressed to gastrointestinal, hepatic, renal, and neurological dysfunction [32].

The pathogenesis of the disease starts when *Rickettsia spp* invades endothelial cells, promoting their dissemination that stimulates cell signaling cascades, which leads to cytokine secretion, infiltration into the vascular cell wall and perivascular space of macrophages, CD4^+^ and CD8^+^ T lymphocytes and NK (Natural Killer) cells (28). As a result of the invasion, the endothelium acquires an activated inflammation phenotype (29). In human *in vitro* models, show that endothelial cells increase the expression and secretion of cytokines IL-1 (interleukin-1), IL-6, and TNFα (tumor necrosis factor α); chemokines such as CXCL8 / IL-8 (interleukin-8), CXCL10 / IP-10 (interferon gamma-induced protein 10), CCL2 / MCP-1 (monocyte chemoattractant protein 1), CXCL9 / MIG (interferon gamma-induced monokine), and CCL5 / RANTES, which lead to the activation and recruitment of leukocytes to the site of infection leading to the potentiation of the inflammatory response and its elimination (28,30–33). Monocytes, immature macrophages that circulate in the bloodstream, once infected promote the spread of the pathogen in the vascular endothelium, causing an increase in microvascular permeability, which is the main pathophysiologic effect in this disease (34–37).

The defense mechanisms against pathogenic rickettsial bacteria in humans are quite complex and poorly understood; nonetheless, there is an understanding supported by evidence from animal models and cell cultures (29). The immune mechanisms by which the host kills and controls rickettsial bacteria are highly dependent on cellular immunity, with a critical role identified for T lymphocytes (38,39). Animal models of rickettsial infections have demonstrated that cell-mediated immunity is essential for complete clearance of these pathogens, in particular CD4^+^ T cells(40); although Walker et al.,2001; Feng and Walker, 2004 and Ismail and Walker, 2005 carried out studies to evaluate the roles of T lymphocyte subsets and revealed that CD8^+^ T lymphocytes also are crucial for the control and killing of these bacteria resistant to phagocytosis. However, current investigation on T lymphocyte subsets in peripheral blood of patients with rickettsial infection is scarce (41,42).

The results and data reported by Escárcega-Ávila et al., 2019, in a serological study carried out on veterinary and administrative personnel report that some of the individuals had been exposed to up to three rickettsial pathogens such as *R. rickettsii, E. canis and A. phagocytophilum* (43), allow us to hypothesize that there is the possibility that these pathogens are involved in human clinical cases. This manuscript aims to follow the disturbance in some immune parameters in patients who presented a clinical scenario of a single rickettsial or ehrlichial infection and coinfection. Surveillance of patients admitted to the Hospital Infantil de Especialidades de Chihuahua with clinical manifestations suggesting a rickettssial infection were analyzed by PCR to detect and identify the presence of *Rickettsia spp, Ehrlichia spp* or *Anaplasma spp*, routine protocol laboratory cabinet tests were performed, and flow cytometry approaches, serum cytokine, and chemokine profiles, and T cells subset were assayed. With this strategy, the presence of co-infections and ehrlichiosis surprisingly were identified and clinical cohorts were contrasted in those considered parameters.

## 2. Materials and Methods

### 2.1. Study subjects and inclusion criteria

The study group included fourteen patients (43% female, 57% male) suspected of rickettsial disease from Children’s Hospital of Specialties of the State of Chihuahua and five healthy controls with ages ranging from 7 to 16 years old, 40% of them were female and 60% male, healthy controls were recruited from the state of Chihuahua, Mexico without a history of a tick bite; autoimmune, cardiovascular, cerebral and / or osteoarticular disorders; acute infectious processes, and ingestion of drugs before sampling. Every child was punctured in the saphenous vein to obtain blood samples collected in EDTA anticoagulant tubes and tubes with cloth activator and gel for serum separation. The patient’s diagnoses required the detection of the gltA gene of *R*. rickettsii and the 16S rRNA gene of *E. canis* by polymerase chain reaction (PCR) using test kits.

### 2.2. Statement of ethics

The study protocol complied with the guidelines established in the Regulations of the General Health Law in matters of health research, the Declaration of Helsinki, and the Good Clinical Practices issued by the National Bioethics Commission. The Research Ethics Committee also approved it of the Faculty of Medicine and Biomedical Sciences of the Autonomous University of Chihuahua with registration number CI-056-19. A written statement that formal consent was obtained from the parent/guardian.

### 2.3. Hematological and serum biochemistry

The blood samples collected were immediately analyzed for the complete hematological and biochemical profiles at Hospital Infantil de Especialidades de Chihuahua – Clinical Laboratory. ALB, albumin; ALT, alanine aminotransferase; AP, alkaline phosphatase; AST, aspartate aminotransferase; Ca, calcium; Cl, chloride; CRE, creatinine; ERY, erythrocytes; DB, direct bilirubin; GGT, γ-glutamyltransferase; GLU, glucose; Hb, hemoglobin; HCT, hematocrit; IB, indirect bilirubin; K, potassium; LEU, leukocytes; LYM, lymphocytes; EOS eosinophils; BAS basophils; MON monocytes; MCH, mean corpuscular hemoglobin; MCV, mean corpuscular volume; Mg, magnesium; MPV, mean platelet volume; Na, sodium; NEU, neutrophils; NR, no results; PLT, platelets; TB, total bilirubin.

### 2.4. DNA isolation, PCR protocol, and Sequencing

The DNA was extracted by column procedure according to the manufacturer’s instructions; then the DNA was quantified using a nanodrop. The molecular diagnosis was performed by end-point PCR test as previously described (44–46). For this purpose, amplifying the 17 kDa protein gene of *Rickettsia* spp., the 16S rRNA ribosomal gene for *Ehrlichia* spp., and *A. phagocytophilum* was performed. Samples positive for the 17 kDa gene were amplified again to amplify the gltA gen to corroborate the genus *R. rickettsia*. In the case of positive samples for *Ehrlichia* spp., nested PCR was performed to identify *E. canis* specifically.

### 2.5. Phylogenetic analysis

Sequences obtained of selected rickettsial species used were identified by BLAST searches of the non-redundant sequences database of the National Center for Biotechnology Information (NCBI). Nucleotide sequences were aligned using the MEGA 11 program (47). A phylogenetic tree was constructed using the maximum likelihood (ML) method in MEGA software. The reliability of the clustering pattern in the phylogenetic tree was tested by bootstrapping, using 1000 pseudo-samples(48,49).

### 2.6. Cytokine and Chemokine Profiles

Cytokine and chemokine levels were evaluated from sera samples obtained from patients with rickettsial disease and control donors. For this, the BD cytometric bead array (CBA) human inflammation kit (Catalog No. 551811), human Th1/Th2/Th17 kit (Catalog No. 560484), and human chemokine kit (Catalog No. 552990) were used following the manufacturer’s instructions. Briefly, the reaction was made from a Master mix containing 5 μL of each antibody. The diluent solution was added to a volume per reaction of 50 μL; then, 50 μL of the test sample and 25 μL of PE were added. The reaction was incubated for 2 h in the dark. Subsequently, 1 mL of wash buffer was added and centrifuged for 5 min at 200× *g*. Finally, 350 μL of wash buffer was added, and read on an Attune NxT cytometer in which 10,000 events were recorded at a flow of 100 μL/min. Results were analyzed with the FlowJo vX.0.7 program. A graph was generated comparing the size (FWD) and the spheres’ complexity (SSC). In this way, the fluorescence of the PE-antibody interest regions was delimited, and the fluorescence intensity corresponding to each cytokine was measured. With these values, the concentration of each cytokine was calculated in picograms per milliliter (pg/mL).

### 2.7. T cell Immunophenotype

T cell subsets were determined with DURAClone Technology (B53328, Beckman Coulter). This panel includes 10 markers in fluorochrome combinations that provide robust population identification, including CD3, CD4, CD8, CD27, CD28, CD45, CD45RA, CD57, CD197 (CCR7), and CD279 (PD-1). Sample preparation and analysis were carried out following the manufacturer’s instructions. Samples were acquired in the Attune NxT cytometer and analyzed in FlowJo software, where the gates strategy to identify the populations was determined (FlowJo version 10; BD Science, San Diego, CA). Gate strategy analysis is shown in Supplementary Figure 1.

### 2.8. Statistical analysis

Data were recorded in a Microsoft Excel spreadsheet and analyzed in the statistical package SAS version 9.0 (SAS Institute Inc., Cary, NC). There were two response variables: 1) the proportion of T lymphocyte and dendritic cell subpopulations and 2) the concentrations of cytokines, chemokines, and soluble proteins of Th’s response, which were dependent on each patient, the clinical findings, and pathogen(s) involved. With these variables, a statistical analysis was performed with the non-parametric Mann-Whitney U and Kruskal-Wallis tests since the data obtained do not comply with the assumptions of normality and homoscedasticity. The Mann-Whitney U test will be used to compare the data of the two groups, healthy and sick children. Follow-up groups will also be performed; these data will be analyzed with the Kruskal-Wallis test that will try to compare the results obtained from the case follow-up samples of the positive patients.

## 3. Results

### 3.1. General view of subjects, molecular diagnosis, and detection of monoinfection and co-infection

Fourteen blood samples were obtained from children positive to rickettsial disease from January to December of 2021. The sex ratio of males to females was eight (57%) to six (43%), respectively. The ages of the individuals oscillated between one to 15 years, with an average of six years (Figure 1A). Six (43%) samples were acquired during March and June, five (35%) in July, and the last three (22%) samples from August to October (one sample each month), Figure 1B. All the samples were analyzed by PCR to detect four (28.5%) patients infected with *R. rickettsii*, six (43%) with *E. canis*, and four (28.5%) with both bacteria in co-infection by yielding amplicons of the expected size for each pathogen, DNA sequencing confirmation was performed, Figure 1C and Supplementary Figure 2, displays PCR results, however none of the samples resulted positive for *A. phagocytophilum*. This report could be the first description of *E. canis* in mono-infection and its participation in co-infection with *R. rickettsii* in pediatric patients.

**Figure 1.**
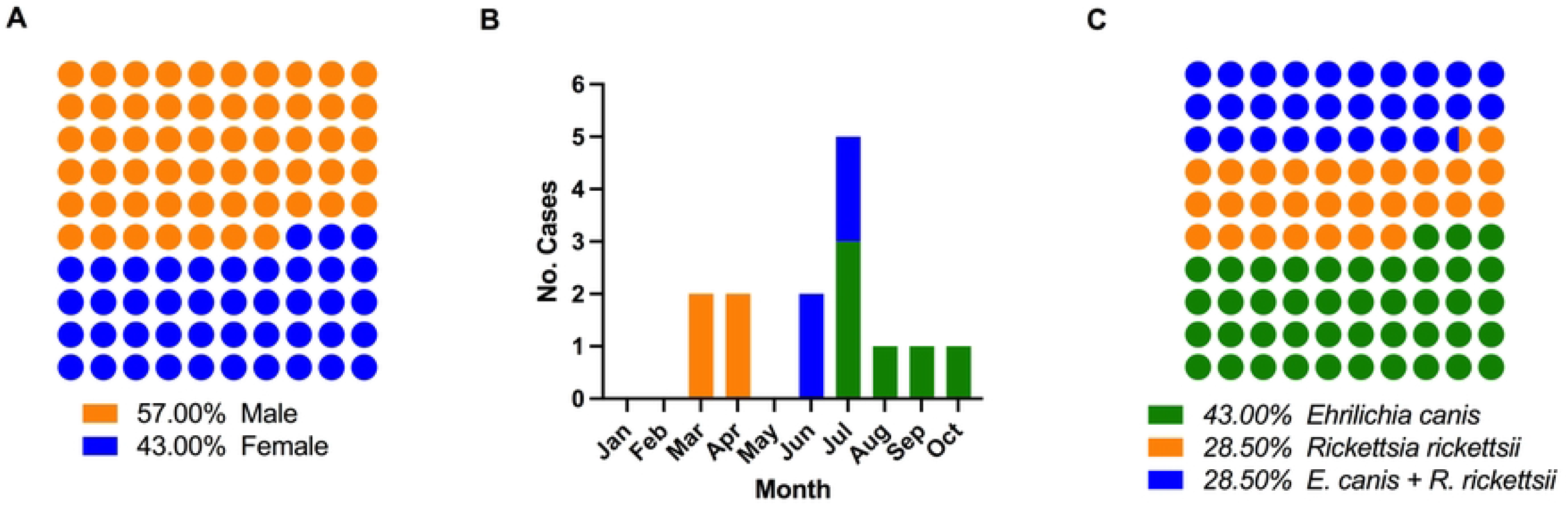
Overview and characteristics from patient samples. A) Gender ratio, B) Case incidence by month for the 2021 year, and C) Ratio of case diseases

### 3.2. Phylogenetic analysis

The results show the efficacy of the 16S and gltA molecular markers in identifying *R. rikettsii* and *E. canis* species. Therefore, it has been shown that these markers are adequate for the phylogenetic discrimination of tick-borne diseases in the State of Chihuahua and could be considered as a pipeline analysis for correctly identifying these pathogens. According to the Bayesian Information Criterion (BIC) score, the best model for the sequences was evaluated based on all the likelihoods. Two phylogenetic trees were generated, one for *R. ricketsii* gltA with the Tamura 3-parameter model (Figure 2A), and the *E. canis* tree (Figure 2B) built with the Kimura 2-parameter model. In both cases, bootstrap values of 1000 bootstrap replicates were used.

**Figure 2.**
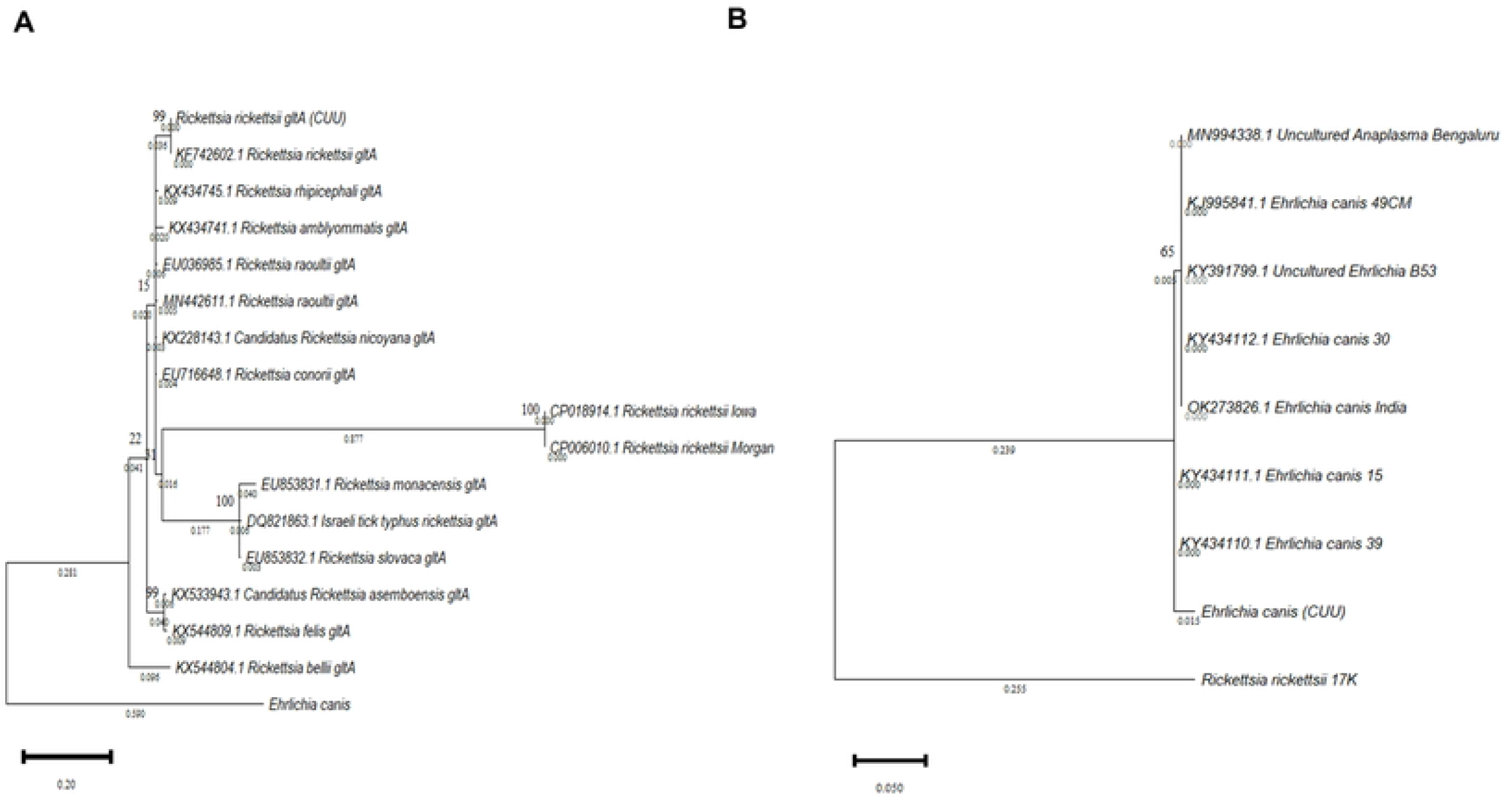
Phylogenetic analysis of sequences obtained from nucleic acid isolated from patient samples. A) Molecular phylogenetic analysis of *Rickettsia* gltA gene, using the maximum likelihood (ML) method generated with the 3-parameter Tamura Model with 1000 bootstrap replicates. B) *Ehrlichia* molecular phylogenetic analysis of the 16S rRNA gene, using the maximum likelihood (ML) method generated with the Kimura 2-parameter model with 1000 bootstrap replicates. Both trees with the highest logarithmic probability are shown. The number of trees in which the associated taxa clustered is shown above the branches. The tree is drawn to scale, with branch lengths below. Each sequence is indicated by its GenBank accession number.

### 3.3. Laboratory cabinet tests

Hematologic and biochemical profiles were recorded from 11 of 14 children’s samples (Table 2. In liver enzymes such as GGT, FA, ALT, AST, and coagulation times (thromboplastin and prothrombin), we found elevated ranges in 100% of the children in the study. 64% of the patients coursed with leukocytosis due to neutrophilia, mainly in positive samples for *R. rickettsii*. 55% of the children presented alteration as hypoalbuminemia, hypoproteinemia, lymphopenia, and thrombocytopenia, at last, normochromic normocytic anemia was observed in 45% of the children. The coinfected group seemed to be the most affected, presenting most the hematologic and biochemical alterations, further data related is available on supplementary Tables 1-3.

**Table 2. Laboratory cabinet tests summary and comparison**

### 3.4. Cytokine and Chemokine Profiles

The cytokines IL-1β, IL-6, IL-8, IL-10, IL-12p70, IL-17, IFNγ, and TNFα were analyzed and compared among the four experimental groups. Significant differences were founded in IL-1β upregulated expression between Rickettsial patients and control donors. A rising tendency was observed in all groups in contrast with control for IL-6 a, with significant output in *Rickettsia* and *Ehrlichia* infected groups. IL-8 and IL-10 cytokines displayed significant differences by all of control and coinfection group for IL-12p70, no significant differences were found; however, a slight increase of IL-17 was recorded in the *Rickettsia* and *Ehrlichia* positive groups in contrast with the control group. Additionally, IFNγ and TNFα laid out significantly increased expression levels between *Rickettsia* and the control group, highlighting the upregulation pattern in TNF samples. Afterward data analysis, the chemokine CCL2/MCP-1 showed no expression difference between the control and *Ehrlichia* positive group and a trend to downregulation in *Rickettsia* and coinfected groups. In CCL5/RANTES, no significative differences were displayed among the control group in contrast with *Rickettsia* and *Ehrlichia*; however, a significantly downregulated expression was found related to the coinfection group. Towards CXCL9/MIG, a significant increase was detected within coinfection and control group by CXCL10/IP-10; significant differences were found in *Rickettsia* and coinfected panels in contrast with control and *Ehrlichia*. Data obtained are shown in Figure 3.

**Figure 3.**
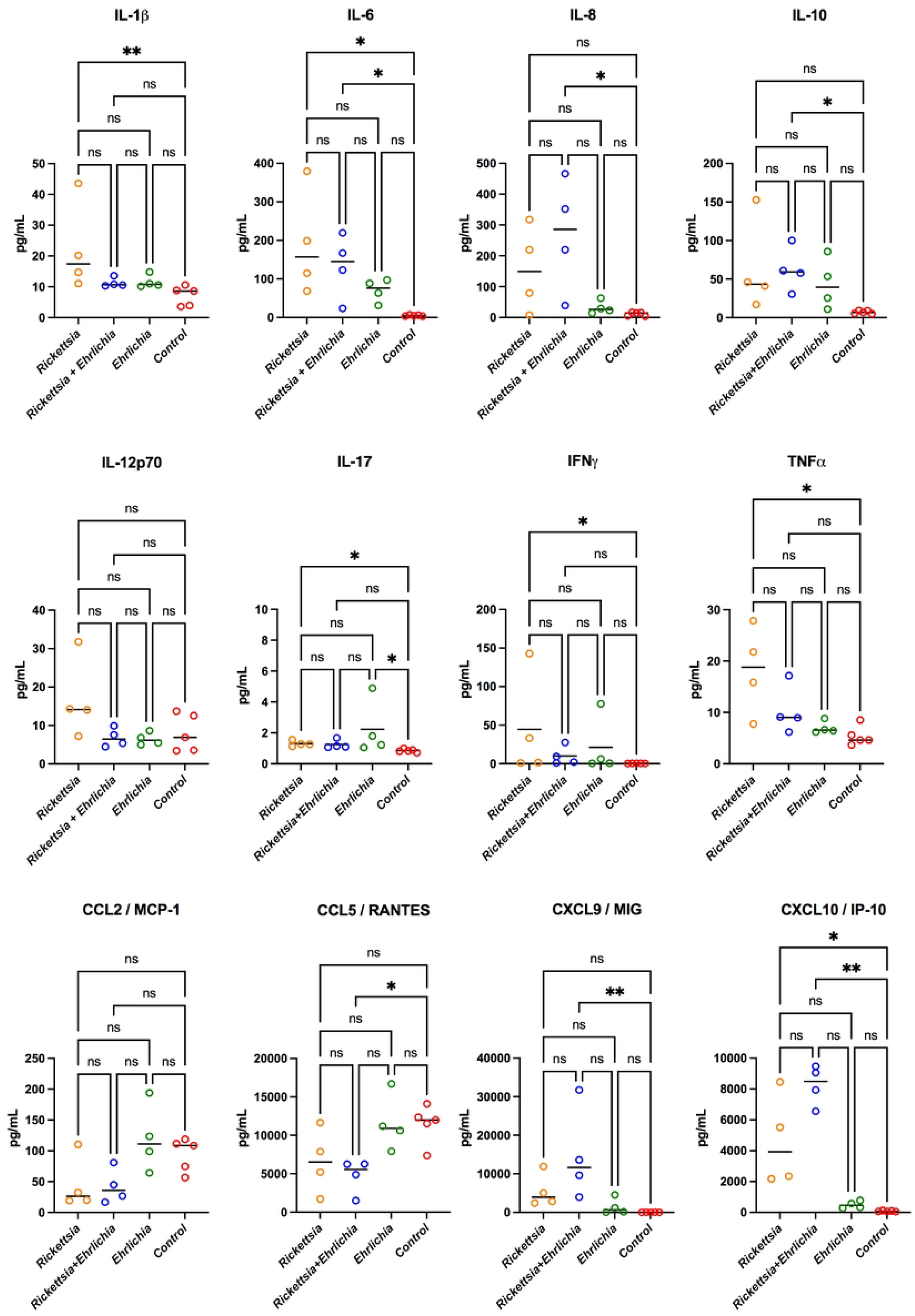
Cytokine and chemokine levels in sera from children with rickettsiosis, coinfection (rickettsiosis and ehrlichiosis), ehrlichiosis, and control. Samples were analyzed in parallel by the bead-based human inflammatory cytokine kit assay. Data are expressed as mean ± SD. Statistical analyzes were performed with the Kruskal-Wallis multiple comparison test. Asterisks indicate differences statistically significant p (* p <0.05, ** p <0.01).

### 3.5. Immunophenotyping CD4^+^ T cells

The analysis of the T cell immunophenotype experiments showed a lower percentage of CD3^+^ T cell population compared to the control group, despite the significant difference in the coinfection group (Kruskal-Wallis, p= 0,0381). Regarding the CD4^+^ T subpopulation cells, non-apparent differences were appreciated; however, a reduction with significant status was detected between *Rickettsia* and *Ehrlichia* groups. In the case of the CD8^+^ T population, the group of *Rickettsia* infection displayed a significative lower proportion of cells in contrast with the control, *Ehrlichia*, and coinfection panels. When T naïve cells CD4^+^ CD45RA^+^CCR7^+^ were evaluated, they displayed a tendency to diminish in contrast with control (no statistical significance). On the other hand, Central Memory (CM) cells and TEMRA showed non-significative difference. Lastly, in Effector Memory (EM) population, it was possible to detect an increase in *Rickettsia* and coinfection groups compared to the control group. Figures 4A and 4B.

**Figure 4.**
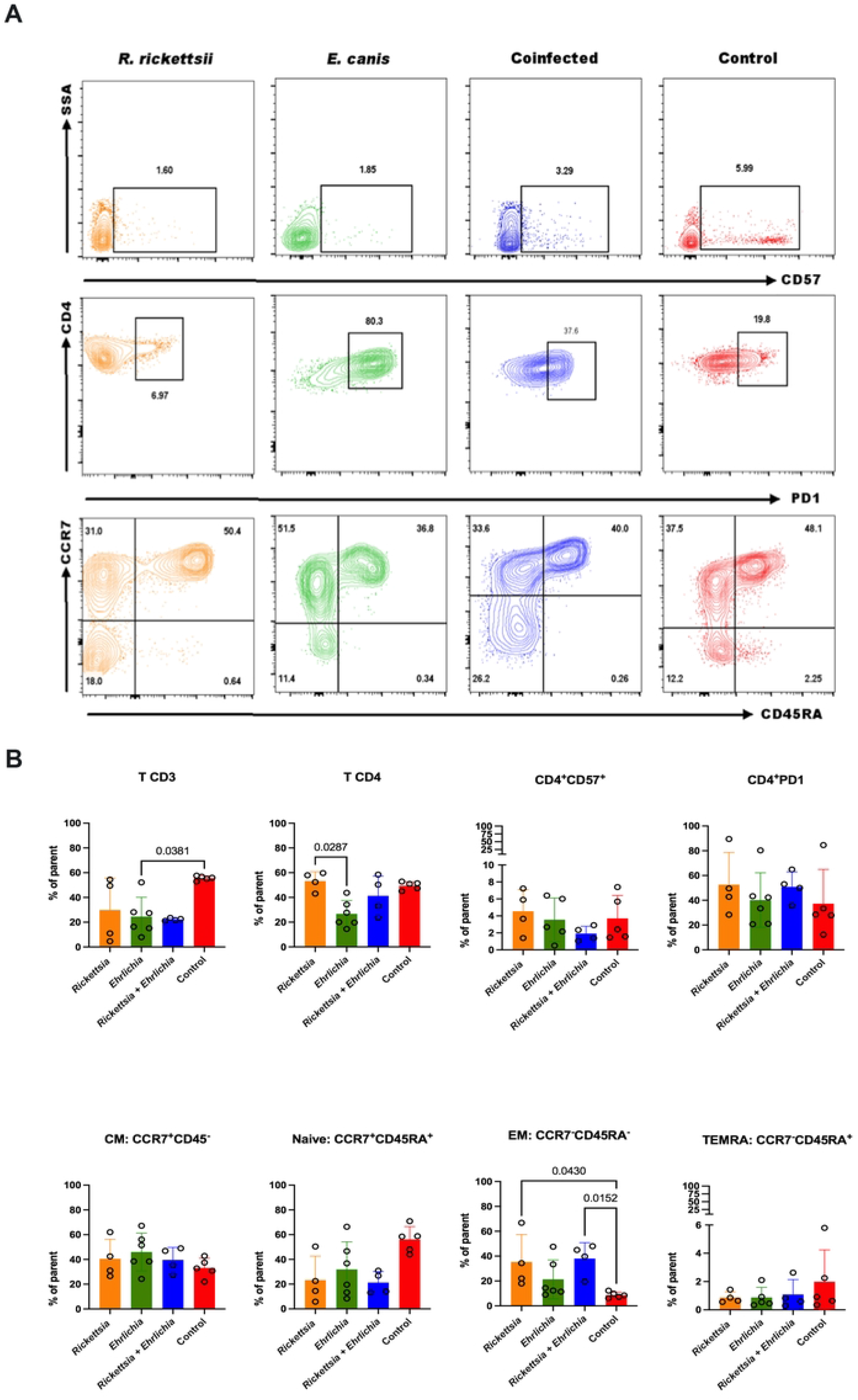
CD4^+^ T cell phenotype analysis from children with rickettsiosis, coinfection (rickettsiosis and ehrlichiosis), ehrlichiosis, and control. Data are expressed as mean ± SD. Statistical analyzes were performed with the Kruskal-Wallis test comparison test. Statistically significant p values are shown in plots.

### 3.6. Immunophenotyping CD8^+^ T cells

For the CD8^+^CD57^+^ T Lymphocytes, the result was significant only between the *Rickettsia* group and the control; however, a slight increase in the populations can be seen in all groups analyzed. In contrast, the CD8^+^PD1^+^ cell population showed significant changes for the coinfected group compared to the control group, also showing increased levels in comparison to the no infection group. In the cell population CD8^+^CCR7^+^CD45^+^ no significant differences were found. In the case of the CD8^+^CCR7^+^CD45RA^-^ population, only a moderately significant increase was detected in the *Ehrlichia* group with respect to the control. Regarding the cell populations CD8^+^CCR7^-^CD45RA^-^, a moderately significant increase of the *Ehrlichia* and coinfected groups with respect to the control was determined, but no significant changes were observed in the other groups. Finally, in the CD8^+^CCR7^-^CD45^+^, cells a tendency to decrease in cells with respect to the control was observed, being significant only in the group of patients infected with *Ehrlichia*. Figures 5A and 5B.

**Figure 5.**
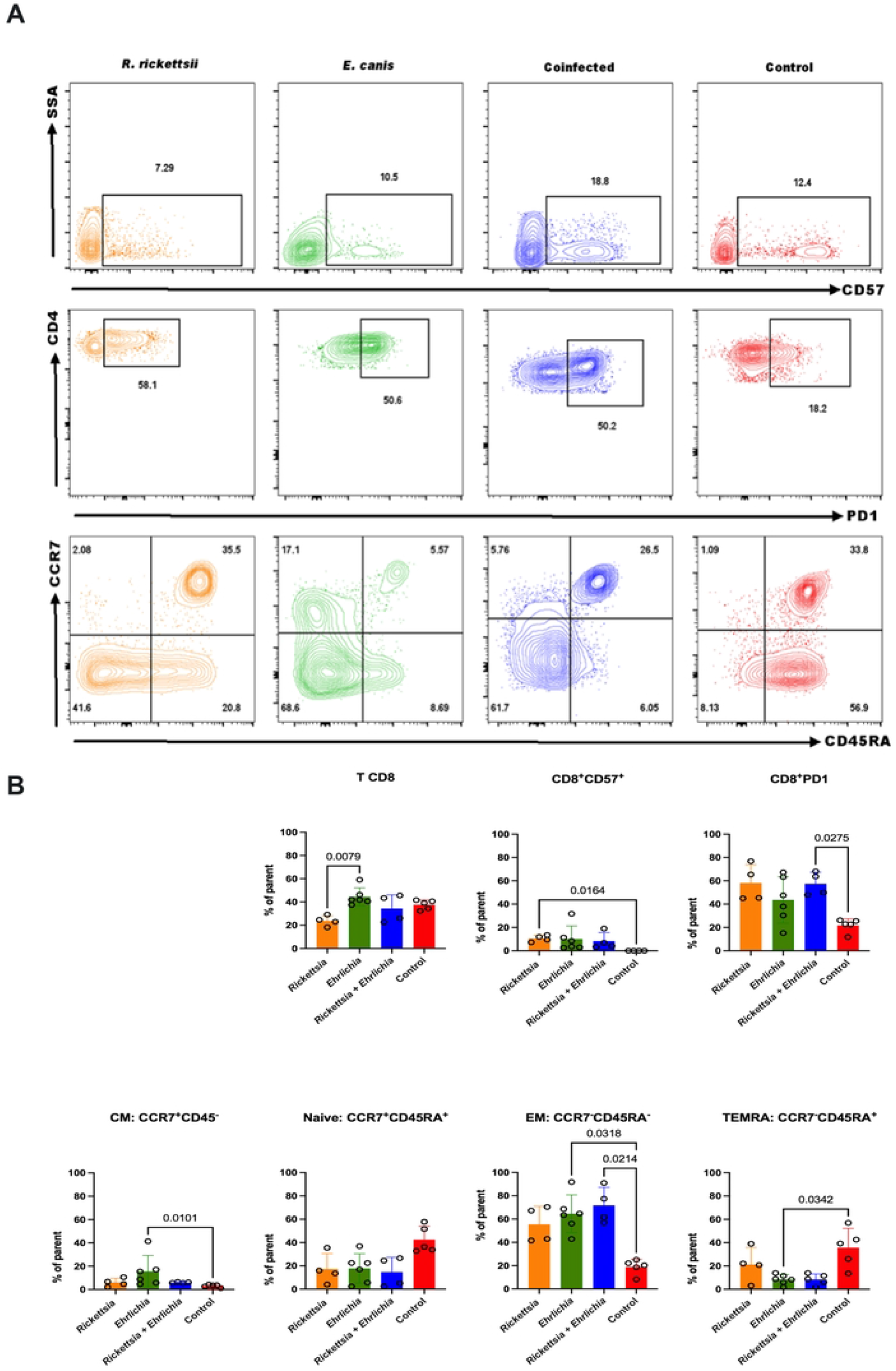
CD8^+^ T cell phenotype analysis in blood obtained from children with rickettsiosis, coinfection (rickettsiosis and ehrlichiosis), ehrlichiosis, and control. Data are expressed as mean ± SD. Statistical analyzes were performed with the Kruskal-Wallis test comparison test. Statistically significant p values are shown in plots.

## 4. Discussion

This manuscript may be the first report of *E. canis* and its association in coinfection with *R. rickettsii* in pediatric patients worldwide. Blood samples were obtained from children suspected of rickettsial infection and were analyzed in 2021; all children positive for these pathogens showed an array of hematological and biochemical clinical alterations. This report provided us the opportunity to work with three different groups of patients with rickettsial diseases, it allowed us to contrast the clinical presentation of infected patients and was possible to determine the cytokine, and chemokine profile in each group, the T cell phenotype, and compare them between groups with the respective control.

Contrasted to the USA, the Northwestern region of Mexico is an endemic zone to *R. sanguineus*, a monotropic vector of dogs, which usually feeds on humans when temperatures are high, and this specie is a competent vector of *R. rickettsia* (50), *A. phagocytophilum* (51), *E. canis* (52), and *A. platys* (53); however, the transmission of other intracellular pathogens (9), *Hepatozoon canis* (54), among others, and their participation in co-infection with these rickettsial diseases, is not excluded.

The association of the acquisition of clinical samples with the scientific infrastructure and reagents to study these valuable samples has been a challenge. In this report, a translational research approach was conducted to provide valuable data to the physicians attending these patients, hoping that the information obtained in the laboratory will be helpful to them to make appropriate clinical decisions and thus provide better care for the patients.

Information about Rickettsial diseases generated in our laboratory has advertised the under-recording of these diseases in Chihuahua and the limited knowledge from the physicians about these zoonotic pathogens. It has been observed that physicians do not contemplate the differential diagnoses and there is an inadequate handling of diagnostic methods for multiple species-specific endemic pathogens leading to under detection of coinfections.

Our results may suggest that coinfections can conduct in a synergic or symbiotic way. Statistically significant differences were obtained for IL-8, RANTES, CXCL9/MIG, and CXCL10/IP-10 chemokines when comparing the coinfected patients and control group, although IP-10 was significant for patients infected by *R. rickettsii* compared to the control group. In cytokine secretion, significant differences were observed for IL-1β, IL-6, IL-17, IFNγ, and TNF-α among the *R. rickettsii* positive group compared to the control group; the coinfected group compared to the control group displayed altered levels of IL-6, IL-8, and IL-10. Finally, significant differences were obtained in all of the subpopulations of CD8+ T lymphocytes when the comparison between the groups positive for *R. rickettsii* and *E. canis* was made.

In agree with other studies, most of the cases in Northern Mexico arise from spring to autumn, with a greater appearance in the summer season, where *R. sanguineus* has highly reproductive activity (52,55). Climate prevailing factors in Chihuahua could relate to the presentation of clinical cases; it is known that at higher temperatures *R. sanguineus* has an aggressive feeding behavior, increasing the risk for bites in humans (56,57). Higher population densities of dogs and *R. sanguineus* have been established as risk factors for acquiring rickettsial diseases in Northern Mexico, as it has been suggested by Escárcega Ávila et al. in a study carried out in Juárez City in 2018, which reported a prevalence in dogs of 43%, 40%, and 28% of *R. rickettsii, E. canis*, and *A. phagocytophilum*, respectively.

Clinical studies reported *E. canis* infection in adults, Perez et al., 2006, detected a prevalence of 30% in patients suspected of rickettsial disease in Venezuela. Silva et al., 2014, reported an isolated case in Oaxaca City from a dog groomer with a subclinical presentation of this illness. Several clinical studies have reported Ehrlichiosis caused by *Ehrlichia chaffensis* around the world. In Mexico, the first registered case of this pathogen was from a 35-year-old man by Góngora et al., 1999 in Mexico State, and the most recent reported case was a homeless man in 2020 who used to coexist with dogs (61).

Coinfections make the diagnosis a challenge; they assert the clinical status of the patient, complicate the treatment, and presumably, in some cases, they work in a symbiotic way or are synergistic (16,17,24). A few cases of coinfections with rickettsial diseases have been reported in Mexico, as reported by Licona-Enríquez et al., 2018, where dengue virus and *R. rickettsii* were diagnosed and responsible for the death of a woman in Sonora State. Dzul-Rosado et al., 2021 reported a clinical case in a child with *R. rickettsii* in coinfection with *Leptospira spp* in Yucatán, with a fatal outcome. In a retrospective study of HIV patients, Paddock et al., 2001 found *E. chaffensis* and *Ehrlichia ewingii* co-infecting these particular hosts located in states belonging to the tick belt in the southeastern region of the United States. Raczniak et al., 2014, described a fatality case in a Native American coinfected with *R. rickettsii* and *Streptococcus pyogenes* in Arizona. Lastly, in North Carolina, Carpenter et al., 1999 carried out a two-year study in which they found patients who presented *R. rickettsii* in coinfection with *E. chaffensis*.

In contrast to results obtained by Rauch et al., 2018 in our study a significant difference was found between the co-infected and mono-infected groups, in which *R. rickettsii* was involved; this could be as a result of the aforementioned, which in some cases of co-infection, the mechanism of action of the pathogens involved boost or act synergistically, increasing the extent of the injury, severity, and complexity of the clinical picture.

Surveillance of Rocky Mountain Spotted Fever (RMSF)is well recorded worldwide (5,18,22,68–70). There are several retrospective, cohort, and case report studies of RMSF in children and adults in northern states like Mexicali (10), Sonora (2,22,25,27,71), Coahuila (4), Yucatán (26,72,73), and Chihuahua (74); however, other Rickettsial pathogens such as *E*.*canis* and *A. phagocytophilum* have not being considered.

Patients with Rickettsial diseases display an unspecific fever syndrome with characteristic temperatures up to 40°C, different clinical picture and several hematological, and biochemical alterations. Likewise, for the patient cohort described in this manuscript, the total of rickettsiosis, ehrlichiosis, and coinfections cases are consistent with previous reports that describe an increase in hepatic enzymes (ALT, AST, and AP), clinical chemical analytes with alterations such as an increased clotting time (CT), prothrombin time (PT) and partial thromboplastin time (PTT) these results are congruent the data reported by Delgado-De la Mora et al., 2018 (27).

The main hematological alteration in patients infected by Rickettsial bacteria is thrombocytopenia; in dogs, this condition is considered as a diagnostic tool for these diseases (75). The literature reports up to 90% of patients with any of these inflammatory diseases present thrombocytopenia and, in some studies, it is related to the severity of the clinical picture in pediatric patients (18,27). Normochromic normocytic anemia and hypoalbuminemia are other alterations similar to those reported by the Centers for Disease Control and Prevention (CDC), 2004 and Zavala-Castro et al., 2008; this alteration is caused by blood loss due to the destruction of the endothelium, damage to liver tissue and physiological compensation mechanisms. It is fundamental to consider that the presentation of the clinical picture and the alterations observed in patients infected with these bacteria will depend on intrinsic host factors such as immune performance, age, and nutritional status, among others (76).

The inflammatory response stimulated by the pathogenic action of rickettsial bacteria triggers immune mechanisms for infection control dependent on cellular immunity and CD4 and CD8 T lymphocytes (77). There are few *in vivo* reports measuring serum pro-inflammatory chemokines and cytokines derived from rickettsial infections in humans caused by *R. rickettsii, E. canis*, and co-infection; in this study, we report the first findings on inflammatory responses during the acute phase of these infections. Significant statistical differences were obtained for the IL-8, RANTES, CXCL9/MIG, and CXCL10/IP-10 chemokines among the coinfected group and the control group; for IP-10 the statiscal difference was significant for the group with *Rickettsia* compared to the control (Figure 2).

Obtained data in this study agrees with data obtained by Rauch et al., 2018; they showed that *R. felis* positive children display a significant difference in the serum levels of IP-10, MCP-1, and IL-8 in a patient cohort compared to the control group; for these cytokines, the authors reported no significant differences in patients coinfected with P. falciparum compared to the positive R. rickettsii group. These chemokines are responsible for attracting leukocytes, T lymphocytes, and NK cells to the infection site to coordinate the elimination and presentation of the pathogen for the activation of T lymphocytes and thus restore homeostasis (21). Valbuena et al., 2003 reported for C3H/HeN mice infected with lethal doses of *R. conorii* a higher expression of chemokines CXCL9 and CXCL10 in the lungs. Besides, Clifton et al., 2005, found an increase in chemokines IL-8 and MCP-1 by measuring mRNA expression in vascular endothelial cells cultured with *R. rickettsii*.

Experiments performed by Rydkina et al., 2005, 2007 in human vascular endothelial cells and umbilical cord cells infected with *R. conorii, R. tiphy*, and *R. rickettsii* described the upregulation of IL-8, MCP-1, IP-10, MCP-2, and RANTES expression. Bechah et al., 2008 obtained upregulated mRNA expression of TNFα, IL1α, IL-6, and CXCL-10, and downregulated expression of CCL-2 and CCL-5 by infecting a line of murine lung microvascular endothelial cells with *R. prowasekii*, thus associating the expression of cytokines in damaged cells with bacterial virulence. The cell endothelium injury caused by these bacteria activates a canonical pro-inflammatory response triggering the expression of cytokines such as IL-1β, TNFα, IFNγ, and IL-6, and an inhibitory stimulus to control this response through IL-10.

Schotthoefer et al., 2017 demonstrated an increase in the serum levels of IFNγ, IL-1β, IL-12p70, and IL-10 in patients with human Anaplasmosis, besides associated the presence in serum of these proteins with the symptomatology, essentially thrombocytopenia, also observed in this infection.

## 5. Conclusion

As previously shown, coinfections raise the fatal outcome scenario and constrain the effective diagnostic window to provide a solid and effective treatment for these patients, who are displayed rickettsial infections. Based on the results obtained, we suggest that coinfections can operate in a synergic or symbiotic way; however, new experiments must be conducted to provide rigorous data about this theory.

Further experiments must be directed to characterize other cell lineages, such as NK cells, dendritic cells, monocytes, and neutrophils, and expand the panel of cytokines. Also, it is essential to address the problem with other types of technologies in a synchronous conjugation of single-cell gene analysis that allows us to generate a more detailed vision of the distortion caused by these pathogens in different cells of the immune system and thus explore the presence of other pathogens that are being left out the simple molecular screening employed.

## Data Availability

All relevant data are within the manuscript and its Supporting Information files.

## Author Contributions

GPES: Conceptualization, project administration, supervision, data analysis, review, and editing of manuscript. LPGR and AEA: conceptualization, data analysis, original draft, review, and editing of manuscript. BEEA: FACS analysis. CAG, phylogenetic analysis MRL, MBA, DMO, EGM: sample collection and patient resources. All authors contributed to this article and approved the submitted version.

## Funding

GPES sincerely acknowledges funding from Consejo Nacional de Ciencia y Tecnología (CONACYT), Ciencia Básica Grant No. A1-S-53789. GPES as part of Laboratorio Nacional de Citometría de Flujo appreciates the support from CONACYT-Apoyos para Acciones de Fortalecimiento, Articulación de Infraestructura y Desarrollo de Proyectos Científicos, Tecnológicos y de Innovación en Laboratorios Nacionales Grant No. 315807. LPR, BEEA, CAG is, 1071642, 790272, 764775 supported from CONACYT, Programa de Becas Nacionales from UACH (Scholarship numbers, respectively).

## Conflict of interest

The authors have no conflicts of interest to declare that are relevant to the content of this article. **Letter is included as a supporting information**

## Notes

### Competing Interest Statement

The authors have declared no competing interest.

### Funding Statement

GPES sincerely acknowledges funding from Consejo Nacional de Ciencia y Tecnologi?a (CONACYT), Ciencia Básica Grant No. A1-S-53789. GPES as part of Laboratorio Nacional de Citometría de Flujo appreciates the support from CONACYT- Apoyos para Acciones de Fortalecimiento, Articulación de Infraestructura y Desarrollo de Proyectos Científicos, Tecnológicos y de Innovación en Laboratorios Nacionales Grant No. 315807. LPR, BEEA, CAG is, 1071642, 790272, 764775 supported from CONACYT, Programa de Becas Nacionales from UACH (Scholarship numbers, respectively).

